# Is 7-days home BP measurement comparable to 24-hours Ambulatory BP Measurement?

**DOI:** 10.1101/2021.10.11.21264844

**Authors:** Sheikh Mohammed Shariful Islam, Chandan Karmakar, Imran Ahmed, Ralph Maddison

**Affiliations:** Deakin University; Virginia Commonwealth University

## Abstract

High blood pressure (BP) or hypertension is a significant risk factor for the global burden of cardiovascular diseases. Home blood pressure measurements (HBPM) have been recommended for hypertension diagnosis, treatment initiation and medication titration, but guidelines for the number of measurements and duration are inconsistent. This study compared the accuracy of 3 home BP measurements per day for seven days with 24-hour ambulatory BP measurements. We examined 24-hour ambulatory BP measurements (ABPM) and HBPM during-morning, afternoon, and evening each day for seven days in healthy community living volunteers. Standardized Bland-Altman scatterplots and limits of agreement (LOA) were used to assess absolute reliability and the variability of measurement biases. We used nonparametric Mann-Whitney U-tests to compare the mean (SD) of the devices. Correlations between HBPM and 24-hour ABPM measurements were statistically significant at p<0.05. The high correlation coefficient (r=0.75) was observed between the systolic BP retrieved from two devices compared to moderate correlation (r=0.46) among diastolic BP. A significant difference was found for systolic BP (P<0.05) between the HBPM and ABPM but was non-significant for diastolic BP (P>0.05). In Bland-Altman plots, the LOA between HBPM and ABPM was 0.07-26.23 mmHg for SBP and 11.24 -16.20 mmHg for DBP. The overall mean difference (bias) in SBP and DBP was 13.08 and 2.48, respectively. Our results suggest that HBPM three times per day for seven days can potentially be used where ABPM is unavailable. Further studies in a diverse group of people with hypertension are needed.

## Introduction

High blood pressure (BP) or hypertension is the leading contributor of cardiovascular morbidity and mortality with increasing prevalence; it accounted for 10.4 million deaths globally in 2017 [1]. A recent campaign screening 1.2 million people in 80 countries found almost 40% of people with hypertension were unaware of their condition, and only 33.2% had their BP controlled [1]. Further, high BP variability is associated with poor cardiovascular outcomes independent of the level of BP [2]. High BP is associated with several adverse health outcomes, including stroke, chronic kidney diseases, diabetes complications, among others [3, 4].

Despite the importance of BP control, lack of timely diagnosis, monitoring and adequate treatment of BP remains challenging worldwide [5]. BP undergoes marked variation over 24-hours and often remains asymptomatic in many people. Therefore, traditional BP measurement at clinics might leave a significant portion of people with potential high BP undiagnosed. Multiple office visits at different timepoints and measurements by expert health care professionals are required to diagnose hypertension, which is impractical for many people. Ambulatory BP measurement (ABPM) is recognized as the reference standard for hypertension detection but is often intrusive, cumbersome for extended use and costlier than current methods [6]. In contrast, home BP measurements (HBPM) performed under a specific protocol using validated equipment can overcome many barriers, such as ruling out white coat hypertension, masked hypertension and record BP changes over time [7].

HBPM is an effective means for detecting hypertension and is more closely associated with hypertension-induced organ damage and adverse cardiovascular outcomes [8]. Several studies have compared HBPM with ABPM and clinic BP, which have highlighted the potential of HBPM as means for hypertension detection and treatment decisions [3, 9-11]. However, there is currently no recommendation for the number of measurements or period of follow-up required each day for HBPM to establish it as a feasible alternative to ABPM. To address this gap, we aimed to determine the comparability of 3 home BP measurements per day at home over seven days with 24-hour ambulatory BP measurement.

## Methods

A convenience sample of 20 healthy adult volunteers was recruited via advertisements at the Deakin University Burwood Campus, Melbourne, Australia. Participants were selected from community settings and attendees at general practice clinics in Melbourne. All participants had normal BP (<130/90 mmHg) at enrollment, were willing to wear an ambulatory BP device for 24-hours and record home BP measurement three times a day for the next seven days. We excluded those taking antihypertensive medications, had severe debilitating diseases, limited mobility, and known mental disorders. The study protocol was approved by the Deakin University Faculty of Health Human Ethics Advisory Group (HEAG-H 135_2017).

We used an ABPM device (A & D Medical Corp, TM-2430) with an adult cuff size to obtain 24-hour BP readings. The ABPM device was programmed to record BP every 30 minutes during 16 hours of that day and every 60 minutes during the remaining 8 hours at night. We provided study participants with a validated home BP device (Omron HEM 7121). They were requested to take BP measurements three times each day during-morning, afternoon, and evening using the provided device and record values using a log sheet. Participants were educated regarding the proper use of each device and how to complete the log sheet. After seven days, all records were collected for data entry and analysis.

The primary outcome of this study was the mean difference between 24-hours ABPM and HBPM measurements for seven days. Data were presented as mean, SD (standard deviation), and range. The mean HBPM measurements were compared against corresponding reference ABPM values. Extreme values were manually investigated to rule out possible measurement errors. As the data did not differ significantly, we used the entire dataset from all devices in these analyses. Pearson product-moment correlation coefficients were estimated. Data were explored graphically using box plots and scatter plots to determine the agreement between the two measurements. Standardized Bland-Altman scatterplots and limits of agreement (LOA) were used to assess absolute reliability and the variability of measurement biases across the measurement range. We used nonparametric Mann-Whitney U tests to compare the mean (SD) of the devices and measure the *P-value* for Box plot analysis. MATLAB 2017a software was used to perform data analyses.

## Results

After seven days of home BP and 24-hour ABPM measurement, we collected 840 and 1530 BP measurements from the respective devices. The majority of participants were young with a mean age of 20.4 (± 5.4) years, highly educated (65% university graduate), employed (60%), and equal participation from both sexes. Mean baseline BP was 112/74 mm Hg. Overall participant characteristics are presented in Table 1.

**Table 1.**
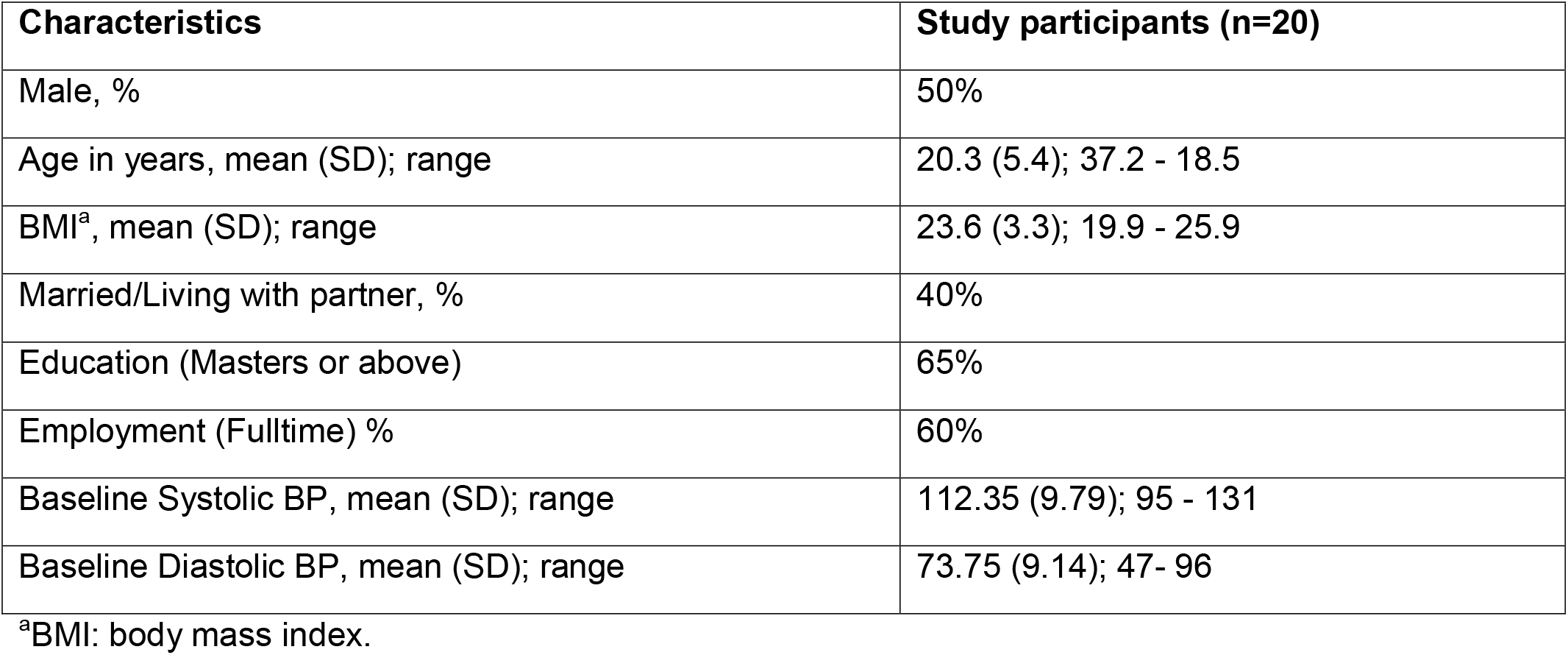
Characteristics of the study participants.

The mean systolic and diastolic values of HBPM and 24-hour ABPM were similar. The correlations between HBPM and 24-hour ABPM measurements were statistically significant at p<0.05. A high Pearson product-moment correlation coefficient (r=0.75) was observed for the systolic BP retrieved from two devices compared to moderate correlation (r=0.46) for diastolic BP (Supplementary Figure 1). A significant difference was found for systolic BP (P<0.05) between the HBPM and ABPM, but this difference was non-significant for diastolic BP (P>0.05).

The Box plot (Figure 1) and the Bland-Altman plots for systolic and diastolic measurements by ABPM and HBPM revealed no systematic biases. LOA between HBPM and ABPM devices was 0.07-26.23 mm Hg for SBP and 11.24 -16.20 mm Hg for DBP. The overall mean difference (bias) in SBP and DBP was 13.08 and 2.48, respectively (Supplementary Figure 2). Overall, for the ABPM device, the mean (SD) systolic and diastolic BP was 125.08 (9.86) mmHg and 73.87 (6.31) mmHg, respectively. For the Home BP device, the mean (SD) systolic and diastolic BP was 112.00 (8.76) mmHg and 71.39 (7.25) mmHg.

**Figure 1:**
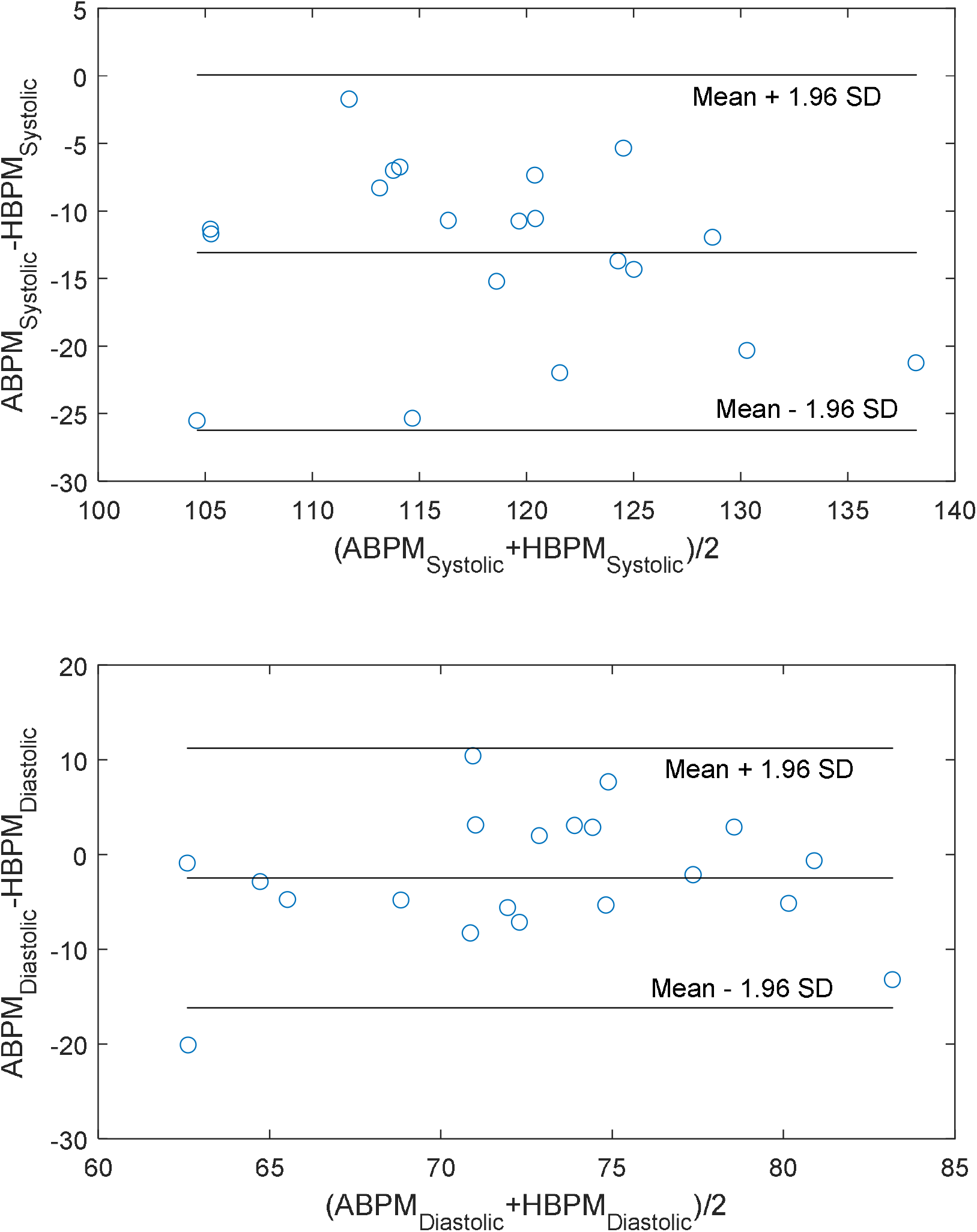
Bland-Altman plots for mean BP measured by ambulatory vs home device.

## Discussion

In this study, we assessed whether a pre-specified home BP measurement strategy provided reliable estimates of BP compared to 24-hours ABPM, the gold standard for detecting hypertension and treatment initiation. Our results suggest that HBPM measurements three times per day for seven days could be an alternative approach to detect hypertension, monitor BP and optimize treatment where ABPM is unavailable or impractical.

BP is an important marker of cardiovascular health and can be measured and monitored by participants in their homes using commercially available HBPM devices. The 2017 American College of Cardiology, the US Preventive Services Task Force and most recent 2020 International Society of Hypertension Global Hypertension Practice Guidelines recommend obtaining out-of-clinic BP measurements to confirm the diagnosis of hypertension before initiating treatment [4, 11, 12]. A study by Bello et al. reported the use of HBPM to estimate home BP reliably and out-of-clinic hypertension diagnosis [8]. The study suggested an average of morning and evening readings of 3 days of HBPM but did not validate the findings against an ABPM. Another study comparing HBPM with automated clinic BP showed that HBPM was much higher compared to clinic BP.

Although HBPM is widely recommended, currently, there is no consensus regarding the recommended number of measurements in a day and follow-up period required to diagnose hypertension compared to ABPM. Previous systematic reviews reported differences in the number of days, readings each day and timings needed to diagnose BP [10, 13]. Bello et al. suggested HBPM using the average of morning and evening readings for three days but compared HBPM with office BP only [8]. Our results correlate with previous studies showing good reliability and agreement between HBPM and ABPM [3, 14, 15]. Other studies have reported negligible or no agreement between ABPM and HBPM [9, 16]. These differences might be due to the different methods of obtaining HBPM measurements, using different devices, population groups, and analysis methods. Our participants were relatively young, employed, and probably had high physical activity compared to previous studies.

HBPM is easy to perform, less burdensome, and cheapest among all BP measurement methods. Our data showed that HBPM performed well compared to ABPM. Current clinic BP measuring techniques are vulnerable to the white coat hypertension phenomena and masked hypertension, resulting in overuse or underuse of medication. In contrast, there are several barriers to the widespread adoption of ABPM in regular clinical practice, specifically in resource-poor settings, rural and remote areas. Therefore, it might be more practical in clinical practise to encourage participants to monitor their BP at home three times per day for a week and report the findings during a consultation. The approach might help clinicians improve hypertension diagnosis and monitor medication effectiveness and titration for optimum BP control. Further, newer internet-enabled HBPM devices enable reporting BP changes more accurately and integrate with clinic record systems providing better BP management insights.

A significant strength of this study was the comparison of HBPM against ABPM in healthy young adults. Our study had few potential limitations. We did not record the exact time point of home BP measurements and activities performed before measurement, which could bias the reports. Participants measured BP three times (morning, evening and night) at their convenience. Our sample size was small, relatively young and non-hypertensive; therefore, the results need to be interpreted with caution. Further research comparing different HBPM devices and approaches in diverse population groups is recommended.

## Conclusion

HBPM measurements three times per day for seven days could be an alternative approach to detect and manage hypertension; however, further studies are recommended in diverse groups with hypertension. This approach could be helpful in low-resource settings where ABPM is not feasible.

## Data Availability

All data in the present study are available online at https://data.jmir.org/2020/1/e22436/authors

## Declaration

Ethics approval and consent to participate: All participants provided written informed consent prior to data collection. The study protocol was approved by the Deakin University Faculty of Health Human Ethics Advisory Group (HEAG-H 135_2017).

### Consent for publication

All authors provided consent for publication.

### Availability of data and materials

Data are available in an open access server [17] (https://data.jmir.org/2020/1/e22436/authors)

### Competing interests

The authors declare no competing interests.

### Funding

SMSI is funded by the National Heart Foundation of Australia and NHMRC Emerging Leadership Fellowship

### Authors’ contributions

Concept: SMSI; Data collection: SMSI; Data analysis: CK; Funding: SMSI; Drafting: SMSI and IA; Critical review: SMSI and RM.

## Acknowledgements

We thank the participants for their valuable time for this research.

## Conflict of interest

## Notes

### Competing Interest Statement

The authors have declared no competing interest.

### Funding Statement

This study was funded by the National Heart Foundation of Australia and NHMRC

### Author Declarations

All participants provided written informed consent prior to data collection. The study protocol was approved by the Deakin University Faculty of Health Human Ethics Advisory Group (HEAG-H 135_2017).

