## Supplementary Figures for "Is 7-days home BP measurement comparable to 24-hours Ambulatory BP Measurement?"

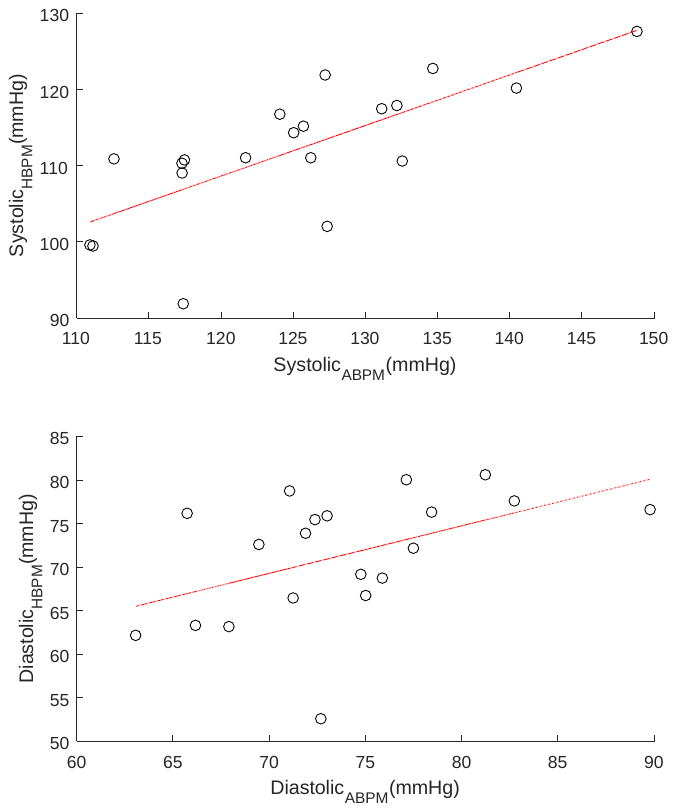

**Supplementary Figure 1:** Correlation between mean ambulatory blood pressure measurement of 1 day and mean home (three times a day) blood pressure measurement of 7 days.

[The top panel shows the correlation between systolic BPs and bottom panel shows the correlation between diastolic BPs. The correlation between systolic BPs is statistically significant (p<0.01) with r=0.75. In contrast, diastolic BPs show lesser correlation with r=0.46 with statistical significance p<0.05.]

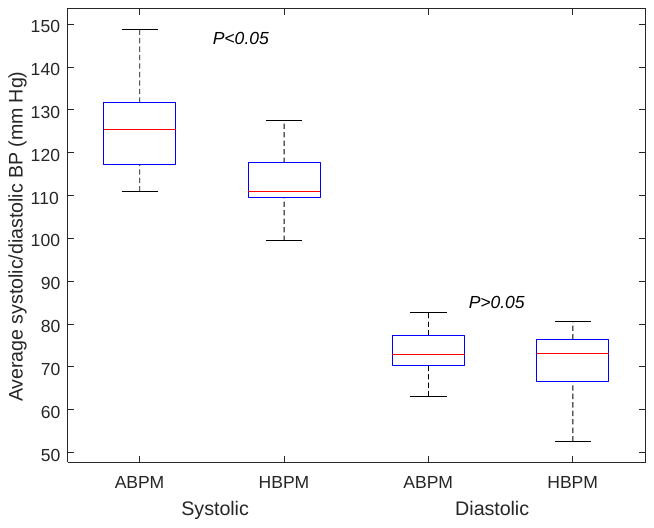

**Supplementary Figure 2:** Box plot showing comparison of average systolic and diastolic BP measured by ambulatory device (Day 1) and home measures over 7 days.

* P-value is calculated using non-parametric Mann-Whitney U Test and P<0.05 was considered indicative of differences between measures.
